# Quantitative susceptibility mapping captures chronic multiple sclerosis rim lesions with greater myelin damage: Comparison with high-pass filtered phase MRI

**DOI:** 10.1101/2021.05.23.21257680

**Authors:** Weiyuan Huang, Elizabeth M. Sweeney, Ulrike W. Kaunzner, Yi Wang, Susan A. Gauthier, Thanh D. Nguyen

**Affiliations:** Department of Radiology, Weill Cornell Medicine, New York, NY, USA; Department of Population Health Sciences, Weill Cornell Medicine, New York, NY, USA; Department of Neurology, Weill Cornell Medicine, New York, NY, USA; Meinig School of Biomedical Engineering, Cornell University, Ithaca, NY, USA; Department of Radiology, Hainan Affiliated Hospital of Hainan Medical University, Haikou, China

## Abstract

**Background:** Chronic active MS lesions with paramagnetic rim can be identified by high-pass filtered (HPF) phase imaging or quantitative susceptibility mapping (QSM).

**Purpose:** The objective was to compare the ability of HPF and QSM to identify MS lesions with greater myelin damage and to distinguish MS patients with increased clinical disability.

**Material and Methods:** Eighty-six patients were scanned with gradient echo sequence for lesion rim detection and FAST-T2 sequence for myelin water fraction (MWF) mapping. Chronic lesions were classified based on the presence/absence of rim on HPF and QSM images (HPF rim+/QSM rim+, HPF rim+/QSM rim-, HPF rim-/QSM rim+, HPF rim-/QSM rim-). A lesion-level linear mixed-effects model with MWF as outcome was used to compare myelin damage among the lesion groups. A multivariate patient-level linear regression model was fit to establish the association between Expanded Disease Status Scale (EDSS) and the number of rim lesions (zero vs. one or more).

**Results:** Of 2229 lesions, 96 (8.8%) were HPF rim+/QSM rim+, 211 (9.5%) were HPF rim+/QSM rim-, and the remainder had no rim. Adjusting for other factors, HPF rim+/QSM rim+ lesions had on average significantly lower MWF than both HPF rim+/QSM rim-(p<0.001) and HPF rim-/QSM rim-(p<0.001) lesions, while the MWF difference between HPF rim+/QSM rim- and HPF rim-/QSM rim-lesions was not statistically significant (p=0.309). Having at least one QSM rim+ lesion was associated with an increase in EDSS compared to having no QSM rim+ lesions, holding all other factors constant (p=0.026). The relationship between having one or more HPF rim+ lesions vs. having no HPF rim+ lesions and EDSS was not statistically significant.

**Conclusion:** QSM identifies chronic MS lesions with paramagnetic rim that on average have greater myelin damage. QSM may be a valuable tool for studying the impact of rim lesions on clinical disability in MS.

## INTRODUCTION

Multiple sclerosis (MS) is a debilitating autoimmune disease of the central nervous system and a leading cause of neurologic disability in young adults (1). MS is characterized by focal demyelination and axonal injury in white matter (WM) as well as chronic progressive neurodegeneration in gray matter (GM) (2). MRI has been widely used to detect and monitor MS lesions for diagnosis and therapeutic management. In the early acute phase, lesions are characterized by active demyelination and breakdown of the blood-brain barrier (BBB) as evidenced by Gadolinium enhancement on post-contrast T1-weighted (T1W) image. Following the transition into the chronic stage after BBB closure, some lesions have the ability to self-repair tissue injury, while others continue to undergo demyelination, which has been attributed to persistent low-grade microglial inflammation behind the sealed BBB (3). An important subset of chronic active lesions have a relatively inactive core surrounded by a slowly expanding rim of continuing demyelination and axonal loss infiltrated by iron-laden activated microglia and macrophages (3-7).

Recently, high-pass filtered (HPF) phase imaging (8-13) and quantitative susceptibility mapping (QSM) (14-17), both derived from the gradient echo (GRE) acquisition, have been developed for identifying rim lesions. This technique has superior sensitivity to rim iron as compared to the conventional T1W and T2-weighted (T2W) sequences. Several studies utilizing either approach have demonstrated that lesions with paramagnetic rim appearance show higher microglial activity (18) and have more tissue damage (19) compared to those without rim, and are associated with worse clinical outcome (20, 21). According to MR physics, both solid and shell lesions may have rim appearance on the HPF phase image depending on lesion geometry, but only shell lesions have rim appearance on the QSM image (22, 23). Therefore, QSM may be more specific than HPF phase for identifying chronic active lesions with greater tissue injury or distinguishing patients with increased disability.

The objective of this in vivo study was to perform a direct comparison of HPF phase imaging and QSM for indicating tissue damage and patient disability. We investigated the association of lesion rim status with myelin damage as measured by myelin water fraction (MWF) (24-26), and we associated the number of rim lesions on HPF phase and QSM images with Expanded Disability Status Scale (EDSS) disability score in MS patients.

## MATERIALS AND METHODS

### Study cohort

This was a cross-sectional retrospective study conducted in a cohort of 85 MS patients (59 women (69.4%), 26 men (30.6%); mean age, 42.5 years ± 10.0 (standard deviation [SD]); range, 26-63 years) selected from an ongoing prospective MS imaging and clinical research database. The database was created under the approval of Weill Cornell Medical College institutional review board and written informed consent was obtained from all participants prior to entry. Consecutive subjects who were imaged between October 24, 2013 and April 19, 2015 and had interpretable brain MRI with multi-echo GRE sequence for QSM and HPF phase imaging as well as Fast Acquisition with Spiral Trajectory and T2prep (FAST-T2) sequence for MWF mapping were included in the study. The final cohort consisted of two patients with clinically isolated syndrome (CIS) and 83 patients with relapsing-remitting MS (RRMS). The mean disease duration was 9.7 years ± 6.0 (range, 2.9-30.0 years) and the mean EDSS was 1.6 ± 1.9 (range, 0.0-7.5; median, 1.5).

### MRI scanning

All patients were imaged on a 3T MR scanner (Signa HDxt; GE Healthcare, Waukesha, WI, USA) using a product 8-channel receiver head coil. The MS brain scanning protocol consisted of 3D T1W BRAVO sequence for anatomical structure, 2D T2W fast spin echo (FSE) and 3D T2W fluid attenuated inversion recovery (FLAIR) CUBE sequences for lesion detection, multi-echo 3D GRE sequence for HPF phase imaging and QSM, 3D FAST-T2 sequence (27) for mapping MWF as marker of demyelination, and gadolinium-enhanced 3D T1W BRAVO sequence for detecting BBB disruption in acute lesions. The typical imaging parameters were as follows: 1) <u>3D sagittal T1W BRAVO</u>: TR/TE/TI = 8.8/3.4/450 ms, flip angle (FA) = 15°, readout bandwidth (rBW) = ±25 kHz, voxel size = 1.2 mm isotropic, parallel imaging acceleration factor (R) = 1.5; 2) <u>2D axial T2W FSE</u>: TR/TE = 5267/86 ms, FA = 90°, rBW = ±50 kHz, echo train length (ETL) = 100, number of signal averages (NEX) = 2, voxel size = 0.6×0.9×3.0 mm^3^ ; 3) <u>3D sagittal T2W FLAIR CUBE</u>: TR/TE/TI = 5000/139/1577 ms, FA = 90°, rBW = ±41.67 kHz, ETL = 162, voxel size = 1.2 mm isotropic, R = 1.6; 3) <u>3D axial multi-echo GRE</u>: TR = 56.9 ms, first TE = 4.3 ms, number of TEs = 10, FA = 15°, rBW = ±62.5 kHz, NEX = 0.75, matrix size = 416×320×48 interpolated to 512×512×48, voxel size = 0.5×0.5×3 mm^3^, scan time = 5.5 min; 4) <u>3D axial FAST-T2</u>: spiral TR/TE = 7.8/0.5 ms, nominal T2prep times = 0 (T2-prep turned off), 7.5, 17.5, 67.5, 147.5, and 307.5 ms, FA = 10°, rBW = ±125 kHz, number of spiral leaves per stack = 32, matrix size = 192×192×32 interpolated to 256×256×32, voxel size = 0.9×0.9×5 mm^3^, scan time = 4 min.

### Image post-processing

Brain QSM was reconstructed from complex GRE image data using the fully automated projection onto dipole fields (PDF) algorithm for background field removal (28) and morphology-enabled dipole inversion with automatic uniform cerebrospinal fluid zero reference (MEDI+0) algorithm for solving the local field-to-source inversion problem (29). HPF phase image was generated by applying high-pass filtering to the complex image acquired at the 4^th^ TE (approximately 20 ms) to remove the confounding effects of the large but smoothly varying background field from the phase image (12). More specifically, a 2D low-pass circularly symmetric Hanning kernel (128×128 voxel) was multiplied with the raw k-space data and Fourier transformed to generate a low-pass filtered complex image. The HPF image was obtained by taking the phase of the complex division between the original unfiltered image and the resulting low-pass image (8). The kernel size was chosen based on the consensus of three readers who visually compared the phase images obtained from two test subjects with kernel size varying between 32 and 192.

Whole-brain MWF maps were computed from FAST-T2 image data using a multivoxel three-pool nonlinear least-squares fitting algorithm with a Laplacian local spatial smoothness constraint (27). The lower and upper T2 bounds for each of the three water pools (in milliseconds) were set to [5 20], [20 200], and [200 2000], which correspond to myelin water, intra- and extracellular water, and free water, respectively (27). MWF was calculated as the ratio of the myelin water signal and the total water signal within a voxel (24).

### Lesion rim classification

Lesions in the brain WM were identified and manually traced on the FLAIR image by an experienced neuroradiologist (W.H., with 11 years of experience) using ITK-SNAP software (version 3.8; http://www.itksnap.org). T2W images were used, if necessary, to break up large confluent lesions on the FLAIR image. Acute Gadolinium-enhancing lesions were excluded from the analysis. The lesion masks were coregistered to the HPF, QSM, and MWF images using the FMRIB’s Linear Image Registration Tool (FLIRT) algorithm and manually adjusted on the MWF image if necessary to account for gross image coregistration error. Lesions with volume smaller than 15 mm^3^ (diameter less than 3 mm in the case of a spherical lesion) were excluded from further analysis in accordance with the currently accepted minimum lesion size on MRI (2). Two expert readers (W.H. and U.K., an MS neurologist with 5 years of experience) independently reviewed lesions on the QSM and HPF images in two separate sessions to minimize recall bias. Lesions were classified on QSM as having a complete or partial hyperintense rim (QSM rim+) relative to the lesion center or without (QSM rim-). Similarly, lesions were identified on the HPF image as having a complete or partial hypointense rim (HPF rim+) relative to the lesion center or without (HPF rim-). In the case of disagreement between the two readers, a third senior MS neurologist (S.G., with 18 years of experience) was called on to determine the final lesion classification. The rim status on the HPF/QSM image, volume on the FLAIR image, and mean MWF value was recorded for each lesion.

### Statistical analysis

Data are presented as mean ± SD. Cohen’s κ was calculated to measure the inter-rater agreement between the first two readers in determining the lesion rim status on the HPF and QSM images. A linear mixed-effects model with mean lesion MWF as an outcome was used to compare the association between lesion type (HPF rim+/QSM rim+, HPF rim-/QSM rim+, HPF rim+/QSM rim-, and HPF rim-/QSM rim-) and lesion myelin damage. The model included a random effect for patients to account for correlation resulting from multiple lesions observed per patients and fixed effects for the lesion type, lesion volume (log-transformed to ensure normality), age, gender, EDSS, disease duration (log-transformed), treatment duration (log-transformed), and MS clinical phenotype (CIS, RRMS). To compare the lesion volume among different lesion types, a linear mixed-effects model with log-transformed lesion volume as outcome and lesion type as a fixed effect was used. Finally, we also fit two separate linear regression models at the patient-level with EDSS as outcome and the number of QSM rim+ lesions (zero vs. one or more) or HPF rim+ lesions (zero vs. one or more) as predictors. Total lesion volume (log-transformed), age, gender, disease duration (log-transformed), treatment duration (log-transformed), and MS phenotype (CIS or RRMS) were adjusted for in these models. The statistical analysis was performed in the R environment (version 3.5.0; R Foundation for Statistical Computing, Vienna, Austria). A p-value of less than 0.05 was considered statistically significant.

## RESULTS

### Descriptive characteristics of rim lesions

A total of 2229 FLAIR-hyperintense non-enhancing lesions were included in the final analysis (Table 1). Of these, 407 (18.3%) lesions could be visualized on the HPF phase image, all of which had rim appearance (HPF rim+), while only 196 lesions (8.8%) had rim appearance on QSM (QSM rim+). All QSM rim+ lesions showed rim appearance on the HPF phase image. Furthermore, all 1822 (81.7%) lesions appearing without rim (HPF rim-) were also found to have no rim on QSM (QSM rim-). Figure 1 shows examples of HPF rim+/QSM rim+, HPF rim+/QSM rim-, and HPF rim-/QSM rim-lesion types. The median volume of HPF rim+/QSM rim+ lesions was 356 mm^3^ (range, 21-6532 mm^3^), which was larger than that of HPF rim+/QSM rim-lesion (median, 177 mm^3^; range, 17-4188 mm^3^) and HPF rim-/QSM rim-lesions (median, 88 mm^3^; range, 15-4270 mm^3^). While 61 (71.8%) patients were found to have at least one HPF rim+ lesion, only 48 (56.5%) patients had at least one QSM rim+ lesion. Among patients with at least one rim lesion, the median of the number of detected HPF rim+ lesions on a per patient basis (median, 5; range, 1-31) was approximately twice that of QSM rim+ lesions (median, 3; range, 1-16).

**Table 1.** Lesion incidence based on the rim appearance on HPF and QSM images of 2229 chronic lesions from 85 MS patients.

|  | **HPF rim+** | **HPF rim-** | **Total** |
| --- | --- | --- | --- |
| **QSM rim+** | 196 (8.8%) | 0 (0%) | 196 (8.8%) |
| **QSM rim-** | 211 (9.5%) | 1822 (81.7%) | 2033 (91.2%) |
| **Total** | 407 (18.3%) | 1822 (81.7%) | 2229 (100%) |

**Figure 1.**
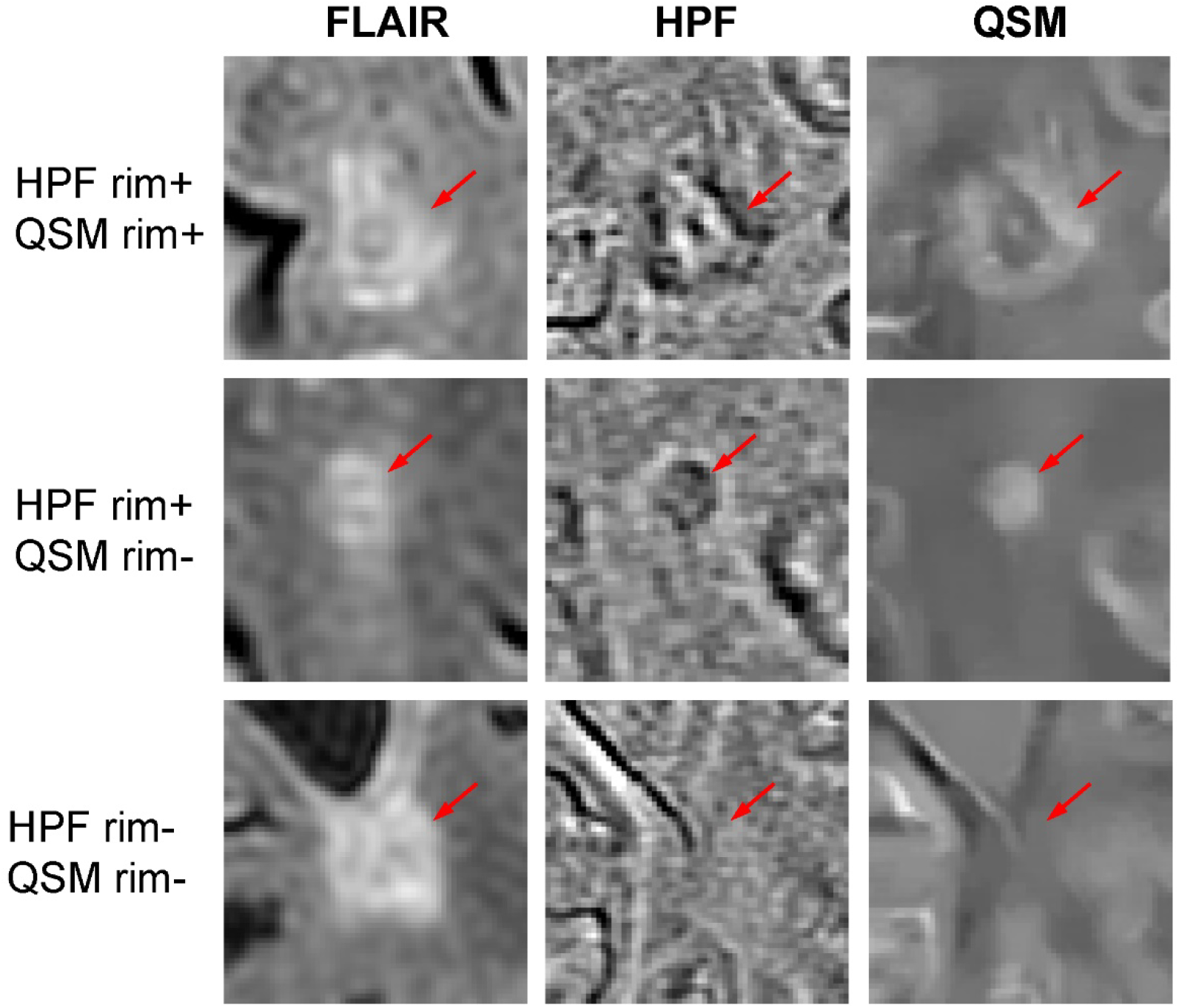
Example of three chronic FLAIR-hyperintense WM lesions with and without paramagnetic rim appearance from three MS patients. Top row: a lesion appears with a dark rim on the HPF phase image and a hyperintense rim on the QSM image, which is consistent with a chronic active lesion with iron-laden microglia and macrophages in the lesion rim. Middle row: a lesion has a rim on the HPF phase image but appears solid without rim on QSM, which may be due to demyelination in the lesion center. Bottom row: a lesion is not visible and has no rim on both HFP and QSM images, suggesting an old chronic lesion with loss of both myelin and iron. In this study, all lesions with rim on QSM also had rim on HPF, and there were no lesions in the HPF rim-/QSM rim+ category.

The inter-rater agreement in calling lesion rim status was found to be moderate on the HPF phase image (Cohen’s κ = 0.43, agreement = 76.1%) and substantial on the QSM image (Cohen’s κ = 0.62, agreement = 93.0%).

### Association between lesion rim status and myelin damage

Figure 2 shows various examples of chronic lesions with paramagnetic rim appearance on both HPF and QSM images (HPF rim+/QSM rim+). These lesions tend to have greater demyelination as measured by MWF imaging compared to QSM rim-lesions including those with rim appearance on the HPF image (Fig.3). Using mean lesion MWF as an outcome variable in the linear mixed-effects model analysis, the presence of lesion rim on QSM (which also signifies rim on HPF, see Table 1) was found to be associated with greater myelin damage, while the presence of rim on HPF but without rim evidence on QSM was not. The only other statistically significant covariate in the model was the log-transformed lesion volume. For every unit increase in log cubic millimeter lesion volume, we found on average a 0.36% decrease in lesion MWF, holding all other factors constant (p<0.001). In our adjusted linear mixed-effects regression model, the MWF of HPF rim+/QSM rim+ lesions on average was 1.08% and 0.93% lower than that of HPF rim-/QSM rim-(p<0.001) and HPF rim+/QSM rim-(p<0.001) lesions, respectively, holding all other factors constant (Fig.4). The observed statistical significance in MWF difference among these lesion groups was maintained when the analysis was repeated for lesions larger than 50, 200, and 500 mm^3^ (corresponding to 1740, 631, and 205 lesions in the statistical samples), which suggests that the result was robust across different lesion sizes. HPF rim+/QSM rim-lesions on average also had lower MWF compared to HPF rim-/QSM rim-lesions; however, this difference was not statistically significant (p=0.309) (Fig.4).

**Figure 2.**
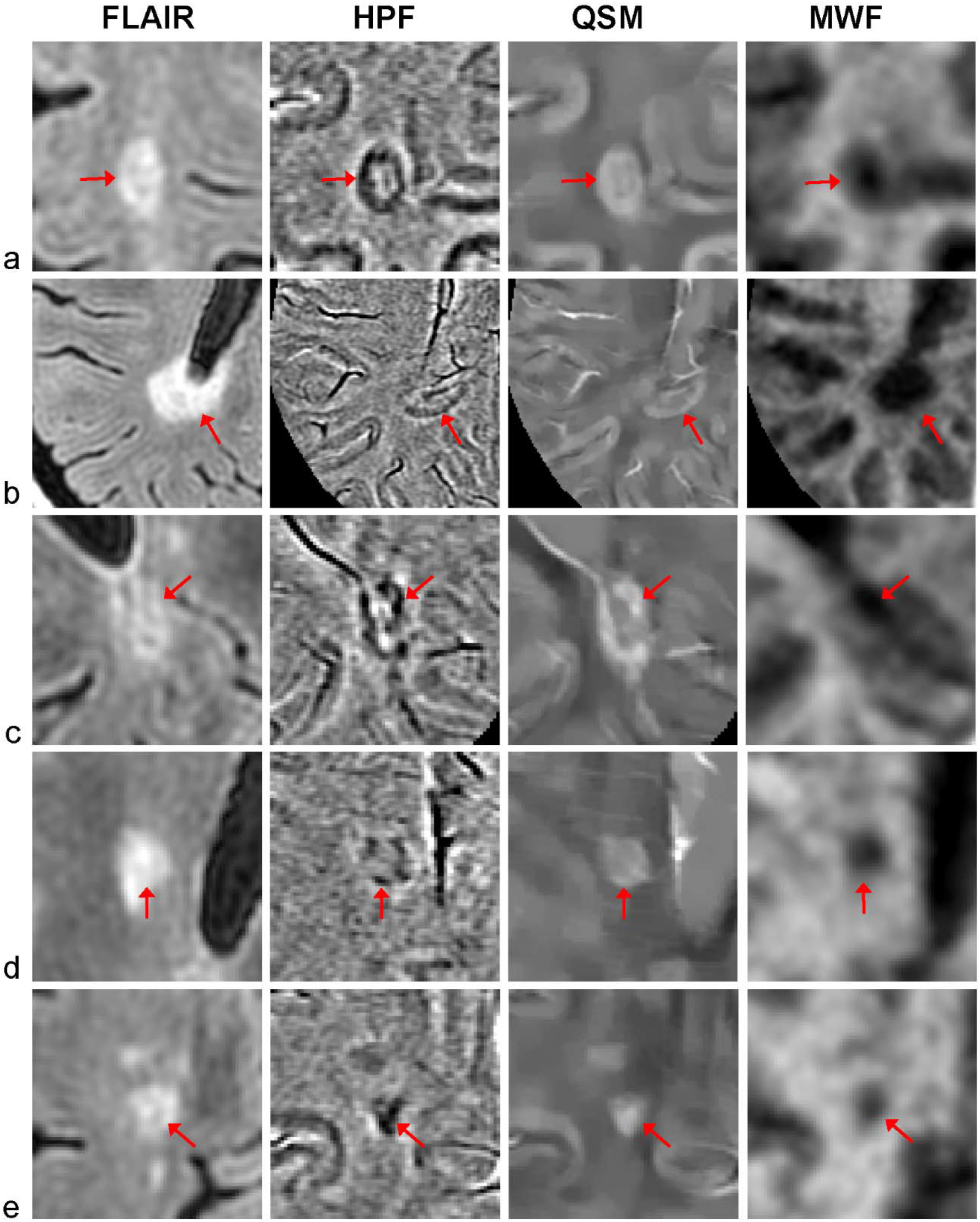
Examples of HPF rim+/QSM rim+ lesions with full or partial rim appearance on both HPF and QSM images and their myelin loss as measured by MWF. Note the improved lesion conspicuity and reduced noise on the QSM image compared to the HPF phase image. The HPF rim+/QSM rim+ lesions tend to have severe demyelination as demonstrated by the much lower MWF value in the lesion center compared to the surrounding NAWM.

**Figure 3.**
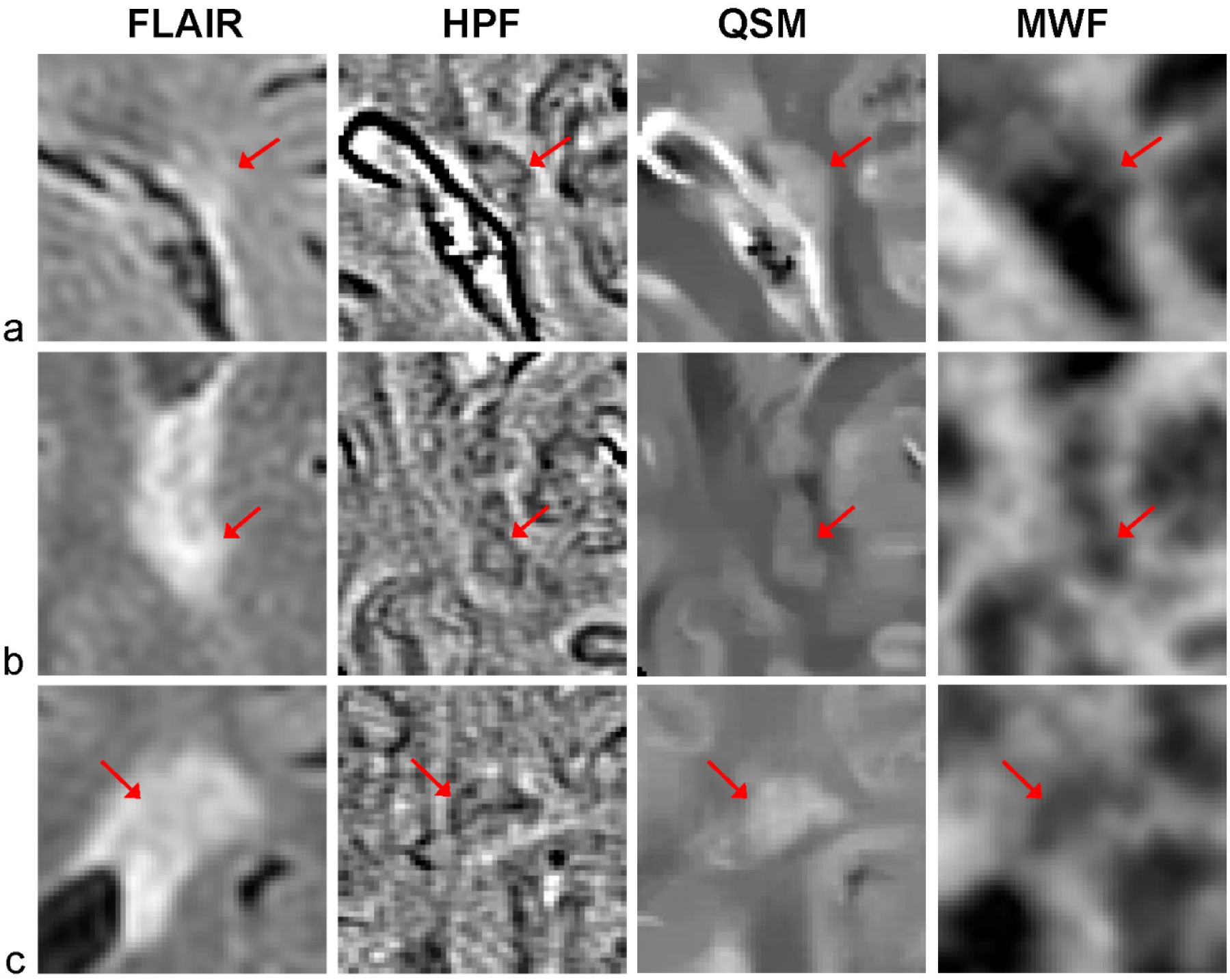
Examples of HPF rim+/QSM rim- lesions with a dark paramagnetic rim on the HPF phase image but no rim appearance on QSM and their MWF values. These lesions show myelin loss, but the degree of demyelination tends to be less severe compared to that of HPF rim+/QSM rim+ lesions shown in Fig.2.

**Figure 4.**
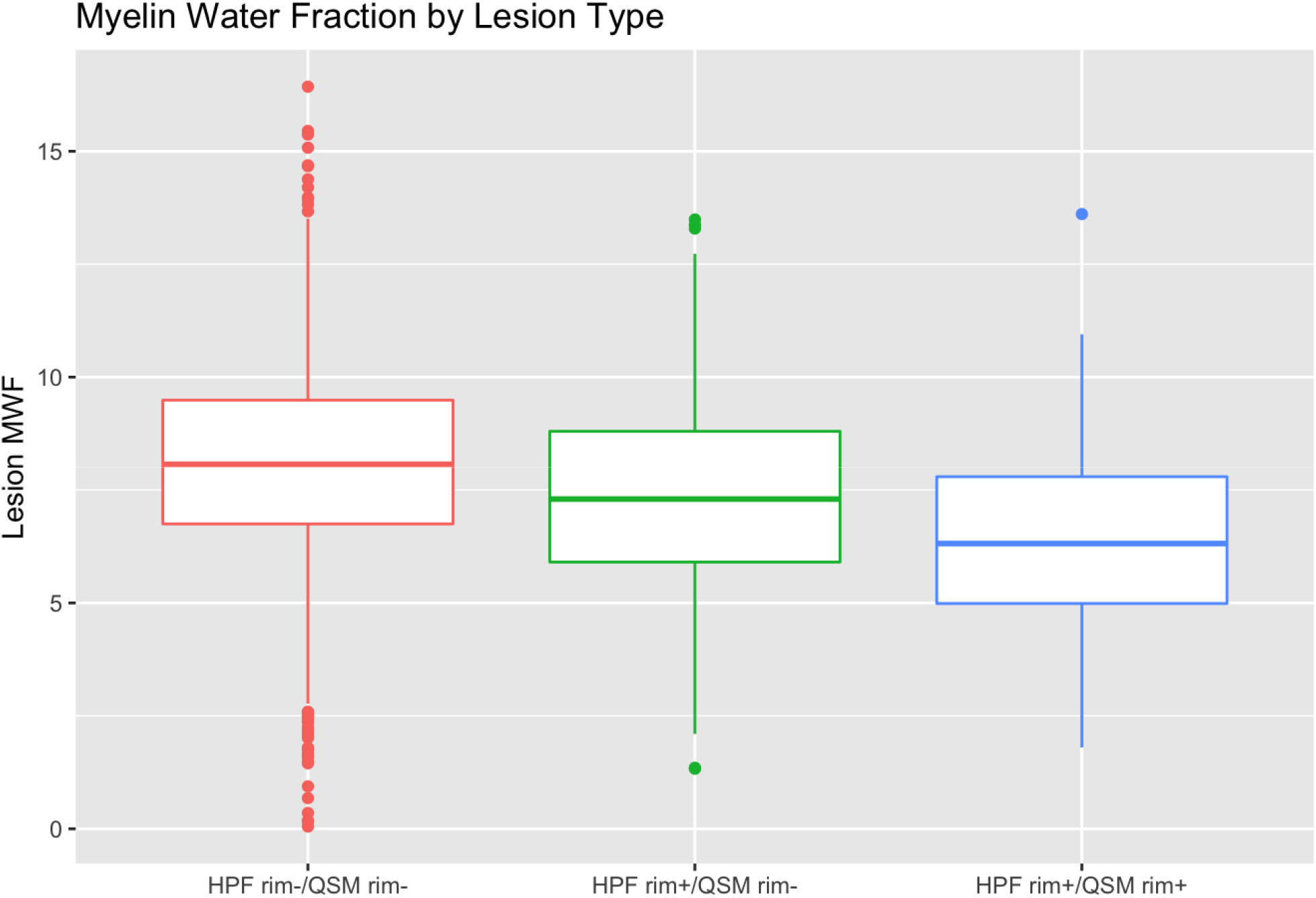
Box plots showing summary statistics of the mean lesion MWF value for the three lesion groups classified based on the paramagnetic rim appearance on both HPF phase and QSM images. Liner mixed-effects model analysis showed that lesions with rim on both HPF and QSM (HPF rim+/QSM rim+) on average have significantly lower MWF compared to the other two groups, holding all other factors constant. Lesions with rim on HPF phase image only (HPF rim+/QSM rim-) tend to have lower MWF compared to those without any rim (HPF rim-/QSM rim-), but the difference did not reach statistical significance.

### Association of lesion rim with lesion volume

Lesions with rim on either QSM or HPF image tend to be larger than those without rim (Fig.5). Using log-transformed FLAIR lesion volume as an outcome in a linear mixed-effects model, lesions with QSM rim (all of which also have HPF rim, see Table 1) were found to be significantly larger than both lesions with rim appearance only on HPF (HPF rim+/QSM rim-) as well as those without rim (HPF rim-/QSM rim-). On average, HPF rim+/QSM rim+ lesions were 1.55 log cubic millimeter larger than HPF rim-/QSM rim-lesions (p<0.001), while HPF rim+/QSM rim-lesions were 0.80 log cubic millimeter larger than HPF rim-/QSM rim-lesions (p<0.001).

### Association between the number of rim lesions and motor disability

In the QSM patient-level linear regression model with EDSS score as an outcome, having one or more QSM rim+ was associated with an average EDSS increase of 1.09 compared to having no QSM rim+ lesions, holding all other factors constant (p=0.026) (Fig.6). The only other variable in the model that was found to be statistically significant was age (p<0.001). For every ten years increase in age, EDSS increases on average by 0.70, holding all other factors constant.

**Figure 5.**
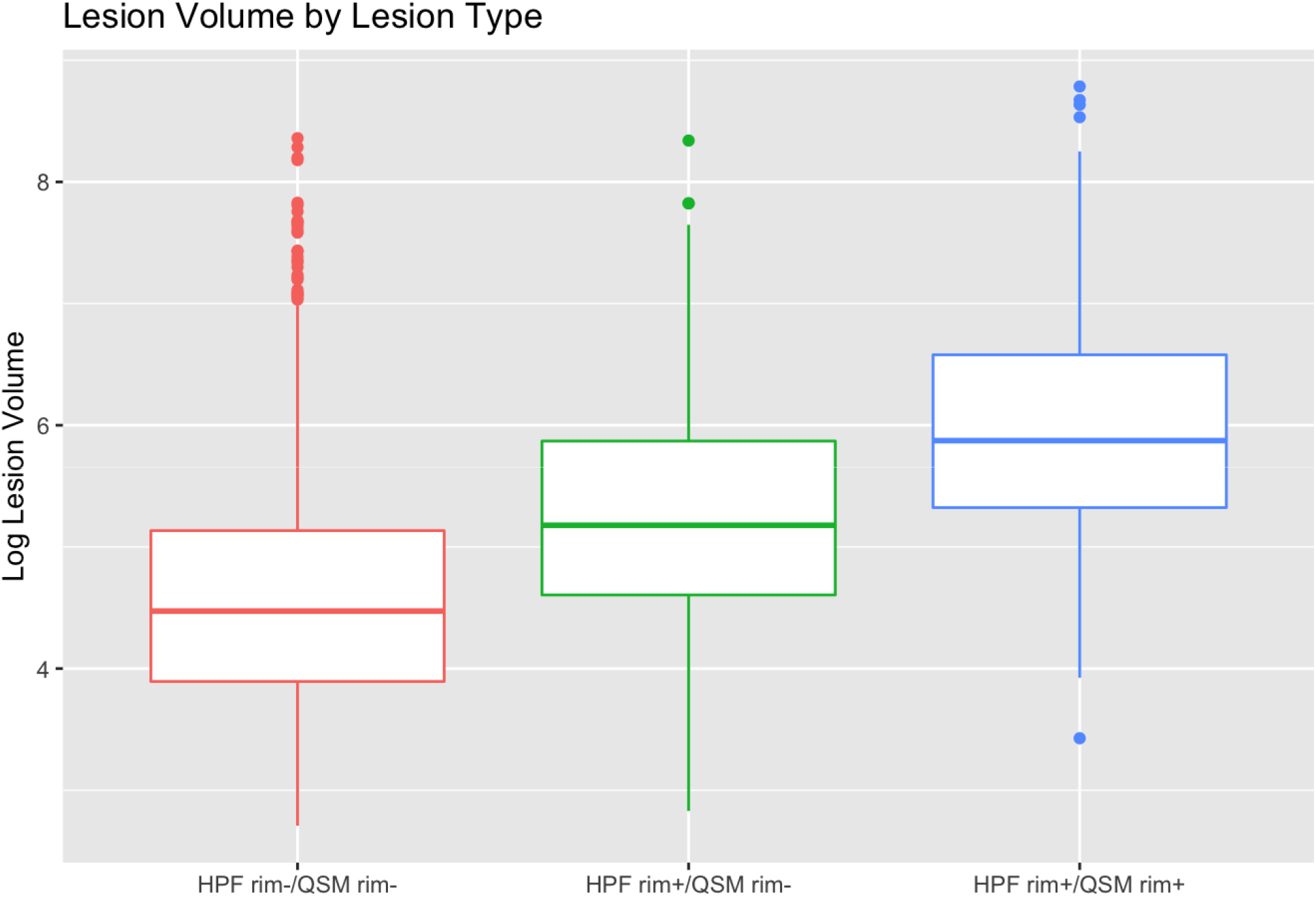
Box plots comparing log-transformed FLAIR lesion volume for three lesion groups classified based on rim appearance on both HPF phase and QSM images. Linear mixed-effects model analysis shows that lesions with rim on either HPF or QSM were on average significantly larger than those without rim. Lesions in the HPF rim+/QSM rim+ group were also significantly larger than those in the HPF rim+/QSM rim-group.

**Figure 6.**
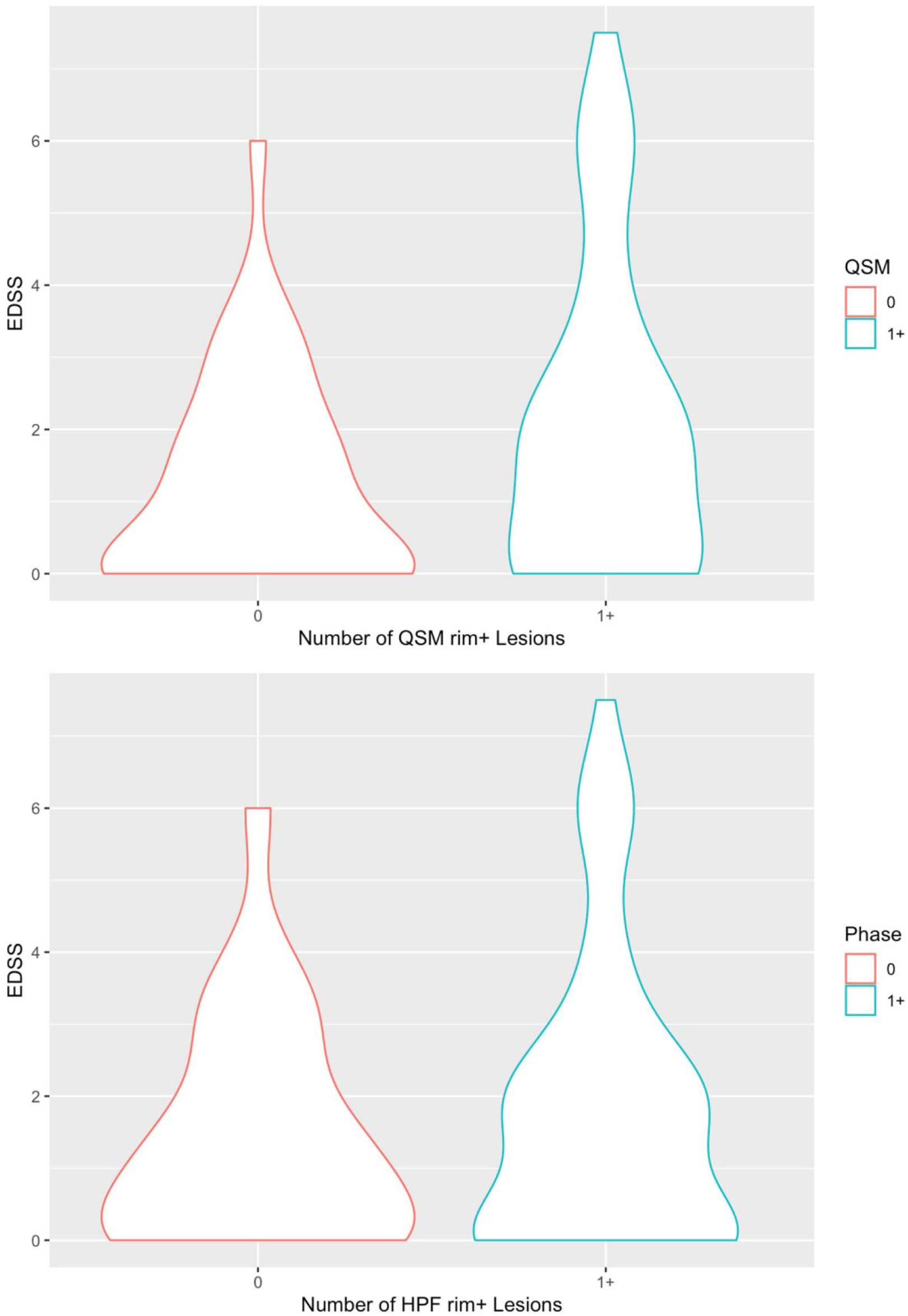
Violin plots showing the distribution of EDSS disability score of MS patients with zero vs. at least one rim lesion on HPF phase (top) or QSM (bottom). Subject-level linear regression model analysis showed that having at least one QSM rim+ lesion is significantly associated with EDSS as an outcome, while having at least one HPF rim+ lesion is not.

In the HPF phase patient-level linear regression model with EDSS as an outcome, having one or more HPF rim+ lesions was not found to be statistically significant versus having no HPF rim+ lesions. Only age was found to be statistically significant in this model (p<0.001). For every ten years increase in age, EDSS increases on average by 0.73, holding all other factors constant.

## DISCUSSION

Data from this HPF phase and QSM comparison study have demonstrated the following findings: 1) On tissue level, lesions with rim on QSM always have rim on HPF (HPF rim+/QSM rim+) and form a distinctive subset of lesions which on average are larger and show greater myelin damage than both lesions with rim only on HPF (HPF rim+/QSM rim-) and lesions without rim (HPF rim-/QSM rim-). While lesions with rim appearance only on HPF (HPF rim+/QSM rim-) also tend to be larger than those without rim (HPF rim-/QSM rim-), the difference in myelin loss between the two groups was not statistically significant. 2) On patient level, patients having at least one QSM rim+ lesion are significantly associated with an increase in EDSS score for motor disability compared with having no QSM rim+ lesion. Patients having one or more HPF rim+ lesion are not statistically significantly associated with an increase in EDSS score compared with having no HPF rim+ lesion. These new findings underscore the added value of QSM compared to the conventional HPF phase imaging in identifying chronic active lesions with more tissue damage and greater impact on the clinical outcome in MS. Our results suggest that QSM may serve as an imaging biomarker of more aggressive chronic active lesions which have been linked to the progressive disease (30).

Both HPF phase and QSM can be derived from the same GRE source data and have shown the presence of iron-positive pro-inflammatory activated microglia and macrophages in the lesion periphery surrounding a relatively inactive demyelinated lesion center (4-6). The paramagnetic rim iron magnetized inside an MRI scanner creates a tissue magnetic field that is the spatial convolution of the iron-related susceptibility distribution with the dipole kernel but dominated by the strong and smooth background field (15). In HPF phase imaging, the background field is removed by HPF, which also removes the low spatial frequency component of the tissue field. Consequently, rim appearance can depend on both the lesion shape and the distribution of other major susceptibility sources such as myelin or veins within and in the vicinity of the lesion (23). For example, a prior phantom study on spherical agarose gel doped with superparamagnetic iron oxide to mimic MS lesions have demonstrated that both solid and shell distribution of iron can give rise to similar phase rim appearance in agreement with theoretical prediction (22, 23). In another study, lesions with histologically confirmed severe demyelination in the lesion core were shown to create a hypointense rim (31). The high-pass filtering effect on the HPF image also tends to accentuate the non-local effect of nearby veins, which have strong susceptibility due to paramagnetic deoxyhemoglobin and can be misinterpreted as lesion rim iron (22). QSM can resolve these signal ambiguities and provide a more truthful depiction of rim iron by directly mapping the susceptibility sources in the brain through spatial deconvolution of the local field (15). Our results in this study have demonstrated that there is a two-fold decrease in the number of QSM rim+ lesions compared to HPF rim+ lesions (406 vs. 196), and that QSM rim+ lesions are associated with increased myelin damage and disability. Therefore, QSM provides a more truthful depiction of lesion susceptibility distribution with improved specificity to lesion rim iron.

There have been a number of investigations of MS lesions with paramagnetic rim appearance on susceptibility-based MR images since the initial reports (9, 10). A recent combined 3T/7T study in 192 patients by Absinta et al (32) found that 56% of subjects had at least one rim lesion on the phase image, which agrees well with the proportion of individuals with at least one rim lesion on QSM (56.5%), but lower than those with at least one rim lesion on HPF (71.8%) in our 3T study. Out of 2229 lesions included in our final analysis (which included all FLAIR-hyperintense lesions with volume of at least 15 mm^3^), the prevalence of lesions with paramagnetic rim was 18.3% on the phase image and 8.8% on QSM, which closely matched the prevalence of 19.9% and 10.1% reported by Li et al in a 7T study of 306 lesions (33). Our rim lesion prevalence on HPF was lower than the 34.7% rate reported by Absinta et al (13) from a 3T study of a limited set of 98 supratentorial chronic lesions from 20 patients. These moderate discrepancies in the detection rate of rim lesions by phase imaging may be attributed to differences in spatial resolution, pulse sequence, main field strength, radiologic criteria for rim definition, or patient characteristics of the study cohort. Harmonization of the GRE data acquisition protocol and standardization of lesion rim definition among research centers will be crucial for establishing rim lesion as a relevant imaging biomarker for MS.

Several studies have analyzed the clinical significance of rim lesions on disease progression and outcome in MS. Harrison et al (20) noted a higher occurrence of rim lesions in more severely disables patients (EDSS≥5) and in those with clinically significant fatigue compared to those with less disability (EDSS<5) or without fatigue. In a study by Absinta et al (32), multivariable analysis found that the presence of 4 or more rim lesions on phase image was independently associated with EDSS score. We also found a significant association with EDSS outcome when only one or more QSM rim+ lesion is present compared to no rim lesions. Our results highlight the potential benefit of QSM over phase imaging for identifying patients with the most aggressive chronic active lesions for targeted treatments. Larger multi-center studies are needed to further confirm this preliminary finding.

Our study has several limitations. First, we used a product GRE sequence with submillimeter in-plane resolution but a relatively thick slice of 3 mm to maintain good signal-to-noise ratio within the scan time of approximately 5 minutes, which may preclude the detection of thin rim in the slice direction due to partial volume effect. In our future work, a QSM acquisition with more isotropic resolution will be optimized and compared with state-of-the-art research pulse sequences for HPF phase imaging such as EPI GRE (13). Second, the acquired spatial resolution of the FAST-T2 sequence (1.2×1.2×5 mm^3^) did not allow reliable estimation of myelin loss in the lesion periphery, which could be a more sensitive marker of the ongoing demyelination in the slowly expanding chronic active lesions (5, 34). Third, we used a cross-sectional design in this study. It would be clinically important to study the relationship between rim lesion evolution (35), including change in rim susceptibility, and tissue damage over time and in response to MS drug treatments. Finally, our study cohort consisted mainly of RRMS patients, and further investigation in MS patients with progressive disease is warranted.

## CONCLUSION

In conclusion, QSM can differentiate chronic MS rim lesions with more severe myelin damage and higher impact on clinical MS disability compared to HPF phase imaging.

## Data Availability

All images used in this work have been de-identified to protect the privacy of human research subjects and are available to interested researchers upon reasonable request.

